# Incretin hypersecretion in gestational diabetes mellitus

**DOI:** 10.1101/2021.11.29.21266983

**Authors:** Louise Fritsche, Martin Heni, Sabine S. Eckstein, Julia Hummel, Anette Schürmann, Hans-Ulrich Häring, Hubert Preißl, Andreas L. Birkenfeld, Andreas Peter, Andreas Fritsche, Robert Wagner

**Author notes:** Corresponding author Louise Fritsche, PhD, University Hospital Tübingen, Department of Internal Medicine IV, Otfried-Müller-Str. 10, 72076 Tübingen, Germany.

## Abstract

**Background:** Incretins are crucial stimulators of insulin secretion after food intake. We investigated the incretin response during an oral glucose tolerance test in pregnant women with and without gestational diabetes.

**Methods:** Subjects underwent a 5-point OGTT with 75 g glucose. We assessed insulin secretion and levels of total GLP-1, GIP, glicentin and glucagon during the OGTT.

**Findings:** We examined 167 women (33 with GDM) during gestational week 26.95 ±2.15. Insulin secretion was significantly lower in women with GDM (P<0.001). Postprandial GLP-1 and GIP were ∼20% higher in women with GDM (all P<0.05) independent from age, BMI and gestational age. GLP-1 increase associated with insulin secretion only in GDM, but not in NGT. Postprandial GLP-1 levels associated negatively with birth weight.

**Interpretation:** The more pronounced GLP-1 increase in women with GDM could be part of a compensatory mechanism counteracting GLP-1 resistance. Higher GLP-1 levels might be protective against fetal overgrowth.

**Funding:** This study was supported in part by a grant from the Federal Ministry of Education and Research (BMBF) (01GI0925) to the German Center for Diabetes Research (DZD).

## Introduction

Gestational diabetes mellitus (GDM) affects 13% of pregnancies with increasing incidence. In pregnancy, insulin resistance develops during the 2^nd^ trimester and is normally compensated by an increase in insulin secretion ^1^. However, if this compensatory increase in insulin secretion falls short, glucose levels rise and GDM develops. When insulin resistance is resolved after delivery, glucose rapidly normalizes. Nevertheless, women who had GDM are at risk to subsequently develop type 2 diabetes ^2^.

After food intake, specialized cells in the gastrointestinal tract release incretin hormones. The best studied incretins are glucagon like peptide 1 (GLP-1) and glucose-dependent insulinotropic peptide (GIP). GLP-1 is cleaved from its precursor proglucagon along with other peptides, including glicentin and oxyntomodulin. Proglucagon gene products in pancreatic α-cells give rise to glucagon.

GLP-1 and GIP are strong enhancers of glucose-stimulated insulin secretion. This incretin effect is reduced in type 2 diabetes and GDM ^3^.

However, only a few studies investigated GLP-1 and GIP concentrations in GDM and reported contrasting results ^4–7^. One study investigated glucagon and detected higher fasting and post-glucose-challenge concentrations in GDM ^8^. No studies on glicentin in pregnancy have been published so far.

### Objective

The aim of our study was to investigate the response of incretins and glucagon during an oral glucose tolerance test in pregnancy in a large cohort of well phenotyped women with normal glucose tolerance and GDM using precise pre-analystics and specific immunoassays.

## Methods

### Subjects

We analyzed data from an ongoing study aiming to characterize metabolic alterations during pregnancy (PREG), a cohort study recruiting women undergoing oral glucose tolerance tests for screening of GDM (NCT04270578). Pregnant women were examined between gestational week 24+0 and 31+6 with a 2h-OGTT with 75 g glucose. GDM was diagnosed using the IADPSG criteria ^9^ and patients were subsequently treated according to national guidelines, which was not part of the study. The detailed study protocol has been described elsewhere ^10^. Informed written consent was obtained from every study participant. The study protocol has been approved by the local ethics boards and the study is conducted in accordance with the declaration of Helsinki. Patients were not involved in the design of the study. Meetings to connect study participants and researchers are held annually.

### OGTT, laboratory analyses and anthropometric assessment

After an overnight fasting period of 12h, all study participants underwent 5-point OGTT with 75 g of glucose. Venous blood was collected at fasting and after 30, 60, 90 and 120 minutes. Plasma glucose and nonesterified fatty acids were measured from sodium-fluoride plasma in an ADVIA chemistry XPT autoanalyzer (Siemens Healthcare Diagnostics) at all timepoints. Serum insulin and C-peptide were analyzed using ADVIA Centaur XPT immunoassay system (Siemens AG) at all timepoints. For the measurement of total GIP, total GLP-1, glicentin and glucagon EDTA-plasma from timepoint 0, 30 and 120 minutes was stabilized with 300 ng/mL of the protease inhibitor aprotinin (Sigma, Merck, Germany) und subsequently processed at 4°C and kept frozen at −80°C until batch measurement. Incretins and glucagon were measured with commercially available ELISA assays (Mercodia, Sweden) according to the manufacturer’s instructions. Height and weight were measured at the day of OGTT. Pregestational weight, parity, gestational age at birth and birthweight were obtained from maternal medical logs. Birth outcome data were available from 135 participants. Insulin sensitivity was calculated using the NEFA-ISI index ^11^. Insulin secretion was assessed with 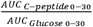 and with ΔC-peptide_0-30_.

### Statistical analysis

All statistical analyses were performed with R Version 3.6.1. In linear regression models skewed variables were log-transformed to approximate normal distribution. Group differences were tested with t-tests for normal distributed variables, Kruskal-Wallis-test for non-normal distributed variables and Chi^2^-test for categorical variables. The provided β coefficients are standardized estimates of the linear regression model terms. A p-value <0.05 was considered statistically significant.

## Results

We analyzed data from 167 women, in 33 GDM was diagnosed. Patients with GDM were older, more likely to be multiparous, but BMI was not different from women with normal glucose tolerance (NGT; Table 1).

**Table 1.** Subject characteristics of the pregnant cohort and birth outcome parameters

| Characteristics | NGT (n = 134) | GDM (n = 33) | p | p adjusted |
| --- | --- | --- | --- | --- |
| Age | 32.71 (4.31) | 35.03 (3.84) | 0.005* | - |
| BMI (kg/m<sup>2</sup>) | 26.90 [23.97, 29.94] | 26.90 [24.72, 30.99] | 0.497 | - |
| Pregestational BMI (kg/m<sup>2</sup>) | 25.37 (5.25) | 25.50 (4.96) | 0.906 | - |
| Gestational weight gain (kg) | 14.06 (5.53) | 11.46 (7.10) | 0.085 | - |
| Parity (%) |  |  |  |  |
| Nulliparous | 69 (51.5) | 9 (27.37) | 0.021* | - |
| Multiparous | 65 (48.5) | 24 (72.7) |  |  |
| Gestational age at OGTT (weeks) | 26.87 (2.16) | 27.27 (2.11) | 0.332 | - |
| Fasting glucose (mmol/L) | 4.28 [4.07, 4.54] | 4.56 [4.33, 4.94] | <0.001* | - |
| 1 h glucose (mmol/L) | 7.70 [6.40, 8.56] | 10.61 [9.56, 11.33] | <0.001* | - |
| 2 h glucose (mmol/L) | 6.11 [5.34, 6.83] | 8.83 [7.72, 9.44] | <0.001* | - |
| Fasting insulin (pmol/L) | 59 [42, 79] | 69[41, 84] | 0.460 | - |
| Fasting C-peptide (pmol/L) | 381.50 [296.50, 496.50] | 466.00 [282.00, 652.00] | 0.150 | - |
| Insulin sensitivity (NEFA-ISI) | 3.34 [2.56, 4.39] | 2.72 [2.01, 4.26] | 0.031* | 0.003*† |
| Insulin secretion (AUC<sub>C-pep 0-30</sub>/AUC<sub>Glucose0-30</sub>) | 179.22 (60.12) | 155.17 (54.53) | 0.037* | <0.001*‡ |
| Birth weight (g) # | 3478.18 (479.84) | 3321.52 (384.38) | 0.084 | 0.24 § |
| Birth length (cm) # | 51.48 (2.37) | 50.48 (2.68) | 0.042 | 0.27 § |
| Gestational age at birth (weeks)<sup>4</sup> | 39.51 (1.59) | 39.16 (1.17) | 0.240 | - |

Data are presented as means (SD), median [IQR] and numbers (%). Group differences were tested with T-test for normal distributed variables, Kruskal-Wallis-test for nonnormal distributed variables and Chi2-test for categorical variables. \*indicates statistical significance. † Adjusted for age, BMI and gestational age, ‡ adjusted for age, BMI, gestational age and insulin sensitivity, § adjusted for gestational age and fetal sex, # n=135. NGT = normal glucose tolerance.

### Glucose and hormones during OGTT

Insulin secretion and sensitivity were lower in GDM (Figure1 A, B). Areas under the curve (AUCs) of glucose and C-peptide were higher in GDM (Figure 1 C, D). While fasting levels of GLP-1 and GIP were similar, the levels at 30min were higher in GDM (Table 2). The AUCs of GLP-1 and GIP were also higher in GDM (all P<0.05, Figure 1 E, F, Table 2). Fasting glicentin, glucagon, and their post-load kinetics were not different between groups (p ≥ 0.1, Figure 1 G, H, Table 2).

**Table 2.** Incretin concentrations during OGTT and AUCs in the pregnant cohort

| Incretin | NGT (n = 134) | GDM (n = 33) | p | p adjusted † |
| --- | --- | --- | --- | --- |
| **GLP-1 min 0**<br>(pmol/L) | 4.48 [3.42, 5.53] | 4.83 [3.56, 6.18] | 0.393 | 0.6 |
| **GLP-1 min 30**<br>(pmol/L) | 9.89 [7.10, 12.83] | 11.31 [8.65, 14.98] | 0.034* | 0.03* |
| **GLP-1 min 120**<br>(pmol/L) | 5.71 [4.55, 8.27] | 6.48 [5.05, 9.30] | 0.175 | 0.3 |
| **AUC GLP-1**<br>(min-pmol/L) | 911.19 [726.59,<br>1207.49] | 1136.24 [913.21,<br>1342.74] | 0.03* | 0.03* |
| **GIP min 0**<br>(pmol/L) | 0.81 [0.81, 1.50] | 0.81 [0.81, 1.76] | 0.312 | 0.5 |
| **GIP min 30**<br>(pmol/L) | 20.21 [14.12, 29.69] | 24.12 [18.76,<br>35.77] | 0.049* | 0.02* |
| **GIP min 120**<br>(pmol/L) | 17.48 [11.92, 23.58] | 19.20 [14.96,<br>27.13] | 0.191 | 0.1 |
| **AUC GIP**<br>(min-pmol/L) | 2022.49 [1421.28,<br>2887.34] | 2532.15 [1899.17,<br>3219.01] | 0.045* | 0.04* |
| **Glicentin min 0**<br>(pmol/L) | 12.37 [8.53, 16.92] | 12.59 [9.56, 16.56] | 0.989 | 0.9 |
| **Glicentin min 30**<br>(pmol/L) | 25.10 [19.26, 34.73] | 28.33 [25.16,<br>35.13] | 0.122 | 0.1 |
| **Glicentin min 120**<br>(pmol/L) | 20.17 [14.97, 30.45] | 21.66 [14.44,<br>29.84] | 0.602 | 0.3 |
| **AUC Glicentin**<br>(min-pmol/L) | 2715.75 [2092.97,<br>3529.69] | 2912.80 [2426.76,<br>3574.03] | 0.198 | 0.1 |
| **Glucagon min 0**<br>(pmol/L) | 1.45 [0.81, 3.42] | 1.43 [0.95, 2.96] | 0.873 | 0.6 |
| **Glucagon min**<br>**30 (pmol/L)** | 1.35 [0.55, 2.20] | 1.15 [0.78, 2.54] | 0.550 | 0.4 |
| **Glucagon min**<br>**120 (pmol/L)** | 0.91 [0.44, 1.53] | 0.74 [0.46, 1.50] | 1.000 | 0.9 |
| **AUC Glucagon**<br>**(min·pmol/L)** | 139.11 [74.68, 236.32] | 124.74 [89.05,<br>262.70] | 0.659 | 0.9 |

Data are presented as median [IQR]. Simple group differences were tested with Kruskal-Wallis-test. † adjusted for age, BMI and gestational week. \*indicates statistical significance.

**Figure 1.**
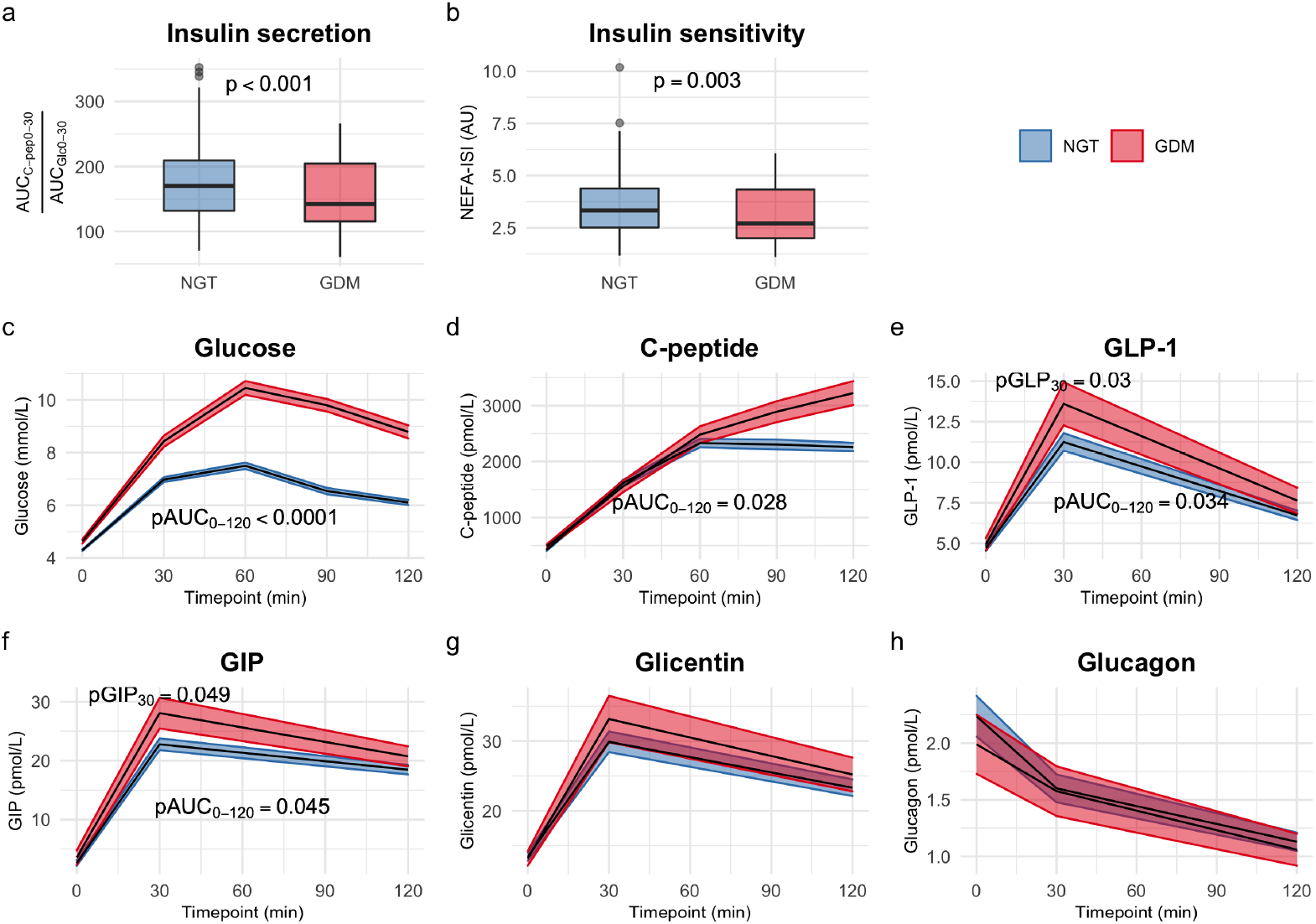
Insulin secretion (A) and insulin sensitivity (B) in women with and without GDM and time course of glucose (C), C-peptide (D), GLP-1 (E), GIP (F), glicentin (G) and glucagon (H) during OGTT. Data are presented as mean ± standard error. Blue represent NGT, red GDM. Group differences between GDM and NGT for AUCs of glucose, insulin and C-peptide were tested with Kruskal-Wallis-test. Differences between groups for insulin sensitivity, incretins and glucagon were tested with multivariate linear regression adjusted for age, BMI and gestational week and for insulin secretion additionally adjusted for insulin sensitivity.

### GLP-1 associates with insulin secretion only in women with GDM

We hypothesized that higher incretin levels in GDM represent a compensatory effect to boost insulin secretion. Therefore, we analyzed the associations of GLP-1 and glucose with insulin secretion (Figure S1). In NGT, increase in glucose between 0-30min associated with insulin secretion (Figure S1 A, P<0.0001). No such association was present for GLP-1 (panel B, p=0.43).

In contrast, women with GDM showed no association of glucose (Figure S1 C, p=0.6) but a positive association of GLP-1 with insulin secretion (panel D, p=0.0004). This remained significant after adjustment for glucose change and basal insulin. Glicentin was similarly associated with insulin secretion in GDM and NGT (Figure S2). For GIP there was no association with insulin secretion (Figure S3).

### Higher GLP-1 associates with lower birth weight

Birth weight negatively associated with AUC GLP-1 (p=0.0182, β=-76.94), 30min GLP-1 (p=0.0248, β=-72.6) and trend for negative association with 30min GIP (p=0.0814, β=-57.45, all models adjusted for gestational age at birth, fetal sex and pregestational BMI). Since GLP-1 and GIP concentrations were higher in GDM, the models were additionally adjusted for GDM. Associations of birth weight with AUC GLP-1 and 30min GLP-1 remained significant (p=0.0296, β=-72.43 and p=0.0381, β=-68.49, respectively).

## Discussion

### Main findings

In this study, we find significantly higher postprandial GLP-1 and GIP levels in pregnant women with gestational diabetes compared to women with normal glucose tolerance, independent from age, body mass index and gestational age. The postprandial GLP-1 increase associated with insulin secretion in women with GDM but not in women with NGT, where insulin secretion was glucose driven. Offspring of women with higher GLP-1 levels also had lower birth weights.

### Interpretation

GLP-1 and GIP have previously been investigated in women with and without GDM in smaller studies with different results ^4,6,7,12^. One study used a different stimulus (mixed meal test) ^6^ and large BMI differences ^6,12^ between NGT and GDM subjects might further confound analyses, because BMI negatively associates with GLP-1 and GIP ^13^. Similar BMI between groups allowed analyzing impact of GDM on incretin responses, without adiposity as confounder.

Intestinal L-cells also secrete glicentin, a potential biomarker of L-cell secretion ^14^. Lack of difference in glicentin between NGT and GDM argues against an unselective L-cell hypersecretion in GDM.

In contrast to findings on glucagon outside of pregnancy ^15^, we and others ^6^ did not detect links to diabetes. This argues against a major contribution of glucagon in the pathogenesis of GDM, while it might have some value in the prognosis of subsequent insulin requirement for the treatment of GDM ^16^.

Of note, women with GDM had lower insulin secretion despite higher GLP-1 concentrations. Our correlational analyses indicate an important contribution of GLP-1 to insulin secretion, especially in GDM. However, this GLP-1 stimulus is still not sufficient to control hyperglycemia. The failure of adequate insulin secretion despite elevated GLP-1 indicates incretin resistance in GDM. The more pronounced GLP-1 response in GDM could be counteracting incretin resistance.

One potential contributor to incretin resistance is genetic background. Polymorphisms in *TCF7L2* associate with elevated GDM risk ^17^ and reduced GLP-1-stimulated insulin secretion ^18^. Another possible contributor might be dipeptidyl peptidase IV (DPP-4) activity, the enzyme degrading GLP-1 and GIP. Liu et al. ^19^ reported no differences of DPP-4 in maternal serum and the unaltered fasting GLP-1 levels in GDM in our study argue against largely different DPP-4 activity. Furthermore, insulin resistance and hyperglycemia which are present in GDM are associated with incretin resistance ^18^.

In our study neonates of women with higher GLP-1 levels during pregnancy had lower birth weight, independent of glycemia. This points towards a protection of GLP-1 against excessive fetal growth. Our findings are in line with a report of negative association between fasting maternal GLP-1 and fetal abdominal circumference and birth weight ^20^. Higher GLP-1, also via decelerated gastric emptying, might reduce postprandial glucose which is associated with fetal overgrowth. If this hypothesis holds true, GLP-1 receptor agonists or DPP-4 inhibitors might have benefits in GDM. To our knowledge, currently no studies test GLP-1R agonists in GDM. One trial reported higher insulin secretion after 16 weeks treatment with the DPP-4 inhibitor sitagliptin in GDM ^21^. However, neither incretins nor birth outcomes were reported.

### Strengths and limitations

The strength of our study is the large number of subjects of our extensively phenotyped cohort of pregnant women. The sample preparation and measurement of incretins was done according to standard operating procedures and with state-of-the-art measuring method. One weakness is the relatively low number of patients with GDM.

## Conclusion

In summary, elevated GLP-1 could be part of a compensatory attempt to counteract GLP-1 resistance in GDM. Higher GLP-1 levels might protect against fetal overgrowth. Our data suggest that not only glucose-stimulated but also incretin-stimulated insulin secretion contributes to GDM. Further studies are needed to translate these findings into improved therapeutic strategies to prevent or treat GDM and circumvent unfavorable impact on the developing child.

## Data Availability

All requests for data and materials will be promptly reviewed by the Data Access Steering Committee to verify whether the request is subject to any intellectual property or confidentiality obligations. Individual-level data may be subject to confidentiality. Any data and materials that can be shared will be released via a Material Transfer Agreement.

## Acknowledgement

We thank all of the volunteers for their participation in the study. We especially thank Ines Wagener, Eva-Maria Stehle, Alexandra Eberle, Dorothee Neuscheler, Henrike Peuker, Elisabeth Metzinger and Heike Runge for their excellent technical assistance.

## Disclosure of interests

RW reports lecture fees from NovoNordisk and travel grants from Eli Lilly. He served on the advisory board of Akcea Therapeutics. In addition to his current work, ALB reports lecture fees from Astra Zeneca, Boehringer Ingelheim, and NovoNordisk. He served on advisory boards of Astra Zeneca, Boehringer Ingelheim and NovoNordisk. Besides his current work, AF reports lecture fees and advisory board membership from Sanofi, Novo Nordisk, Eli Lilly, and AstraZeneca. In addition to is current work, MH reports research grants from Boehringer Ingelheim and Sanofi (both to the University Hospital of Tuebingen) and lecture fees from Sanofi, Novo Nordisk, Eli Lilly and Merck Sharp Dohme. None of the other authors report a conflict of interest directly related to the content of this work.

## Contribution of authorship

LF researched and analyzed data and drafted the manuscript. JH, SN, and RW researched data. HUH, ALB, HP, AS, and AF contributed to the discussion and interpretation of the results. AP supervised the laboratory measurements and interpreted results. RW and MH supervised the project. All authors contributed to discussion and approved the final manuscript prior to submission. RW is the guarantor of this work and, as such, had full access to all the data in the study and takes responsibility for the integrity of the data and the accuracy of the data analysis.

## Details of Ethics Approval

The study protocol has been approved by the ethics board of the University of Tuebingen (No. 218/2012BO2, 06/15/2012).

## Funding

This study was supported in part by a grant from the Federal Ministry of Education and Research (BMBF) (01GI0925) to the German Center for Diabetes Research (DZD).

## Registration

The PREG Study is registered with clinicaltrials.gov NCT 04270578.

## Abbreviations

AUC: Area under the curve
DPP-4: Dipeptidyl Peptidase IV
GDM: Gestational diabetes mellitus
GIP: Glucose dependent insulinotropic peptide
GLP-1: Glucagon like peptide-1
GLP-1R: Glucagon like peptide-1 receptor
IGI: Insulinogenic index
NEFA-ISI: Nonesterified fatty acids - Insulin sensitivity index
NGT: Normal glucose tolerance
OGTT: Oral glucose tolerance test

## Notes

### Author Declarations

Ethics board of the University of Tuebingen gave ethical approval for this work (Protocal number 218/2012BO2, 06/15/2012)

