## Supplementary material for "Incretin hypersecretion in gestational diabetes mellitus": Figure S1, Figure S2, Figure S3

Short running title: Incretins in GDM

Louise Fritsche[1,2], Martin Heni[1,2,3,4], Sabine S. Eckstein[1,2], Julia Hummel[1,2], Anette Schürmann[2,5], Hans-Ulrich Häring[1,2,3] Hubert Preißl[1,2], Andreas L. Birkenfeld[1,2,3], Andreas Peter[1,2,5], Andreas Fritsche[1,2,3], Robert Wagner[1,2,3]

1. Institute for Diabetes Research and Metabolic Diseases of the Helmholtz Center Munich at the University of Tübingen, Tübingen, Germany
2. German Center for Diabetes Research (DZD), Neuherberg, Germany
3. Department of Internal Medicine, Division of Endocrinology, Diabetology and Nephrology, Eberhard Karls University Tübingen, Tübingen, Germany
4. Institute for Clinical Chemistry and Pathobiochemistry, Department for Diagnostic Laboratory Medicine, University Hospital Tübingen, Tübingen, Germany.
5. German Institute of Human Nutrition, Potsdam-Rehbrücke, Germany

Table of contents
Figure S1……………………………………………………………………………….. 2
Figure S2……………………………………………………………………………. 3
Figure S3……………………………………………………………………………. 4

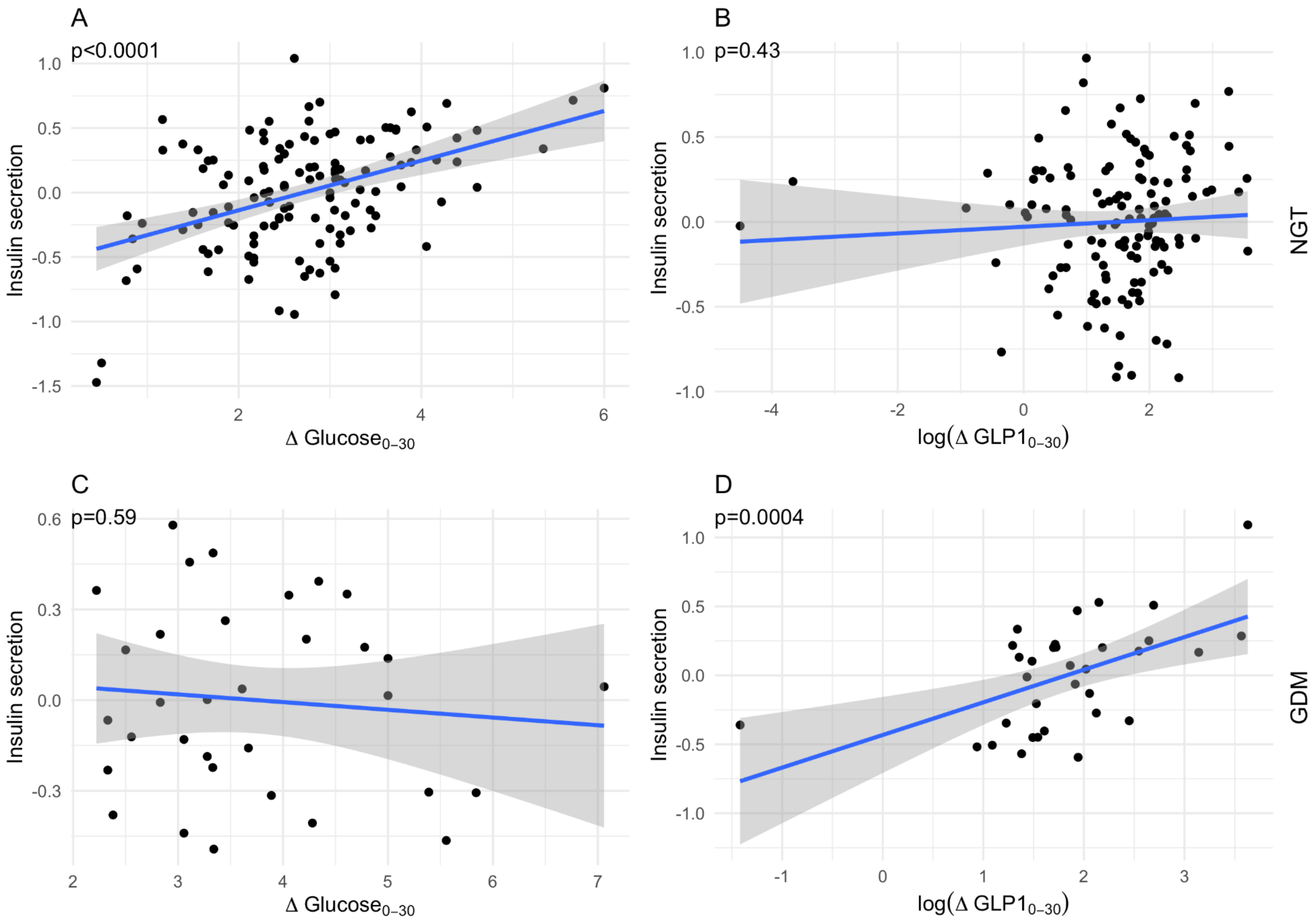


**Figure S1** Association of GLP1 and glucose increase from minute 0 to 30 during OGTT with insulin secretion ($\Delta C - Peptide_{0-30}$) in pregnant women with normal glucose tolerance (NGT) (A, B) and in women with GDM (C, D). Variables were residualized for BMI, basal GLP1, basal insulin, basal glucose and $\Delta GLP1_{0-30}$ (panel A and C) and $\Delta Glucose_{0-30}$ (panel B and D) respectively.

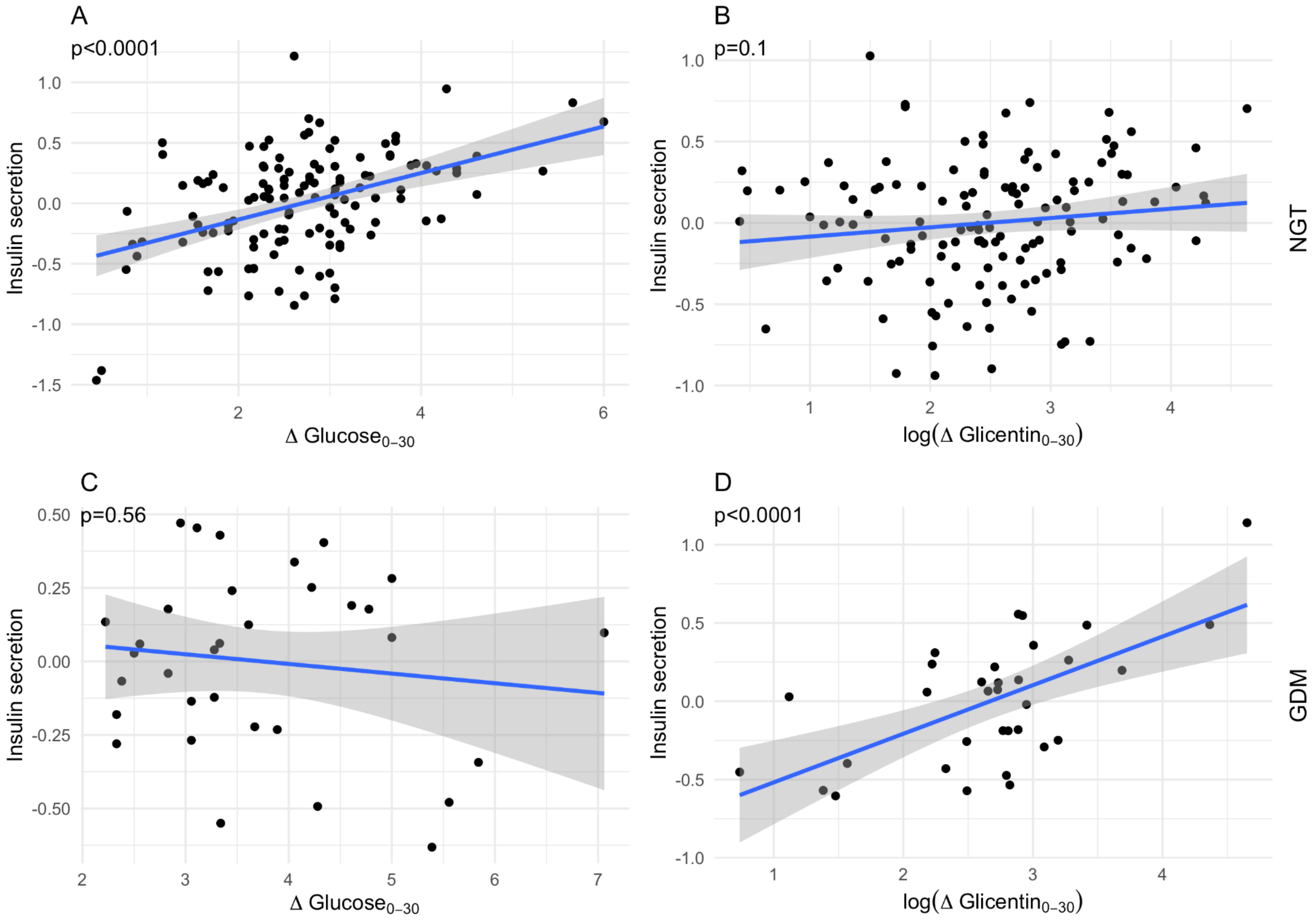


**Figure S2** Association of glicentin and glucose increase from minute 0 to 30 during OGTT with insulin secretion ($\Delta C - Peptide_{0-30}$) in pregnant women with normal glucose tolerance (NGT) (A, B) and in women with GDM (C, D). Variables were residualized for BMI, basal glicentin, basal insulin, basal glucose and $\Delta\text{Glicentin}_{0-30}$ (panel A and C) and $\Delta Glucose_{0-30}$ (panel B and D) respectively.

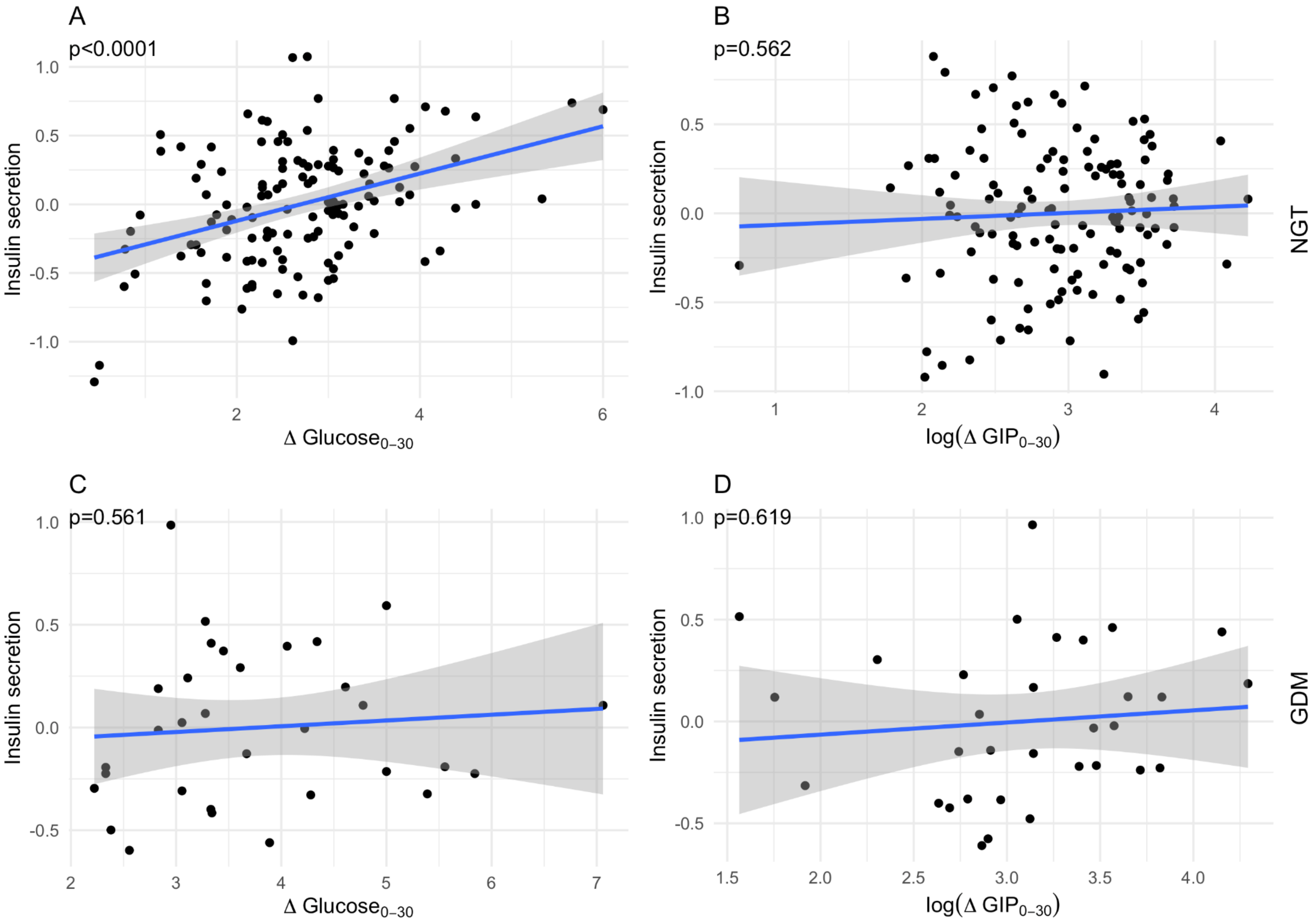


**Figure S3** Association of GIP and glucose increase from minute 0 to 30 during OGTT with insulin secretion ($\Delta C - Peptide_{0-30}$) in pregnant women with normal glucose tolerance (NGT) (A, B) and in women with GDM (C, D). Variables were residualized for BMI, basal GIP, basal insulin, basal glucose and $\Delta \mathrm{GIP}_{0-30}$ (panel A and C) and $\Delta Glucose_{0-30}$ (panel B and D) respectively.
